# Fat-enlarged Axillary Lymph Nodes are Associated with Node-Positive Breast Cancer in Obese Patients

**DOI:** 10.1101/2021.02.17.20246504

**Authors:** Roberta M. diFlorio-Alexander, Qingyuan Song, Dennis Dwan, Judith A. Austin-Strohbehn, Kristen E. Muller, William B. Kinlaw, Todd A. MacKenzie, Margaret R. Karagas, Saeed Hassanpour

**Affiliations:** Department of Radiology, Dartmouth-Hitchcock Medical Center, 1 Medical Center Drive, Lebanon, NH 03756; Department of Biomedical Data Science, Dartmouth College, 1 Medical Center Drive, Lebanon, NH 03756; Department of Internal Medicine, 2100 Dorchester Ave, Carney Hospital, Dorchester, MA 02124; Department of Pathology, Dartmouth-Hitchcock Medical Center, 1 Medical Center Drive, Lebanon, NH 03756; Department of Medicine, Dartmouth-Hitchcock Medical Center, 1 Medical Center Drive, Lebanon, NH 03756; Department of Epidemiology, Dartmouth College, 1 Medical Center Drive, Lebanon, NH 03756; Department of Computer Science, Dartmouth College, Hanover, NH 03755, USA

## Abstract

**Purpose:** Obesity-associated fat infiltration of organ systems is accompanied by organ dysfunction and poor cancer outcomes. Obese women demonstrate variable degrees of fat infiltration of axillary lymph nodes (LNs), and they are at increased risk for node-positive breast cancer. However, the relationship between enlarged axillary nodes and axillary metastases has not been investigated. The purpose of this study is to evaluate the association between axillary metastases and fat-enlarged axillary nodes visualized on mammograms and breast MRI in obese women with a diagnosis of invasive breast cancer.

**Methods:** This retrospective case-control study included 431 patients with histologically confirmed invasive breast cancer. The primary analysis of this study included 306 patients with pre-operative MRI and body mass index (BMI) > 30 (201 node-positive cases and 105 randomly selected node-negative controls) diagnosed with invasive breast cancer diagnosed between April 1, 2011, and March 1, 2020. The largest visible LN was measured in the axilla contralateral to the known breast cancer on breast MRI. Multivariate logistic regression models were used to assess the association between node-positive status and LN size adjusting for age, BMI, tumor size, tumor grade, tumor subtype, and lymphovascular invasion.

**Results:** A strong likelihood of node-positive breast cancer was observed among obese women with fat-expanded lymph nodes (adjusted OR for the 4^th^ vs. 1^st^ quartile for contralateral LN size on MRI: 9.70; 95% CI: 4.26, 23.50; p < 0.001). The receiver operating characteristic curve for size of fat-enlarged nodes in the contralateral axilla identified on breast MRI had an area under the curve of 0.72 for predicting node-positive breast cancer and this increased to 0.80 when combined with patient and tumor characteristics.

**Conclusion:** Fat expansion of axillary lymph nodes was associated with a high likelihood of axillary metastases in obese women with invasive breast cancer independent of BMI and tumor characteristics.

## Introduction

Obesity affects more than 30% of adult women, and obese women have an increased risk of breast cancer with increased risk of axillary node metastases and up to 68% increased breast cancer-specific mortality compared to normal-weight women [1, 2]. Decreased survival is experienced by both pre-menopausal and post-menopausal obese women; and studies have reported no appreciable differences in screening patterns among obese women and normal-weight women suggesting that biologic factors are likely responsible for higher breast cancer risk and mortality [2–5]. While the exact mechanisms accounting for the poor prognosis of breast cancer in obese women are not fully understood, increased available estrogen, insulin, and tumor-promoting characteristics of dysregulated obese adipose tissue have been proposed as contributing factors [1, 6, 7]. Axillary lymph node (LN) status is one of the most important independent prognostic indicators of survival, with reports of up to 14% decrease in 5-year survival associated with a single metastatic axillary node [8, 9]. These findings suggest that targeting nodal metastases in breast cancer treatment may significantly impact breast cancer mortality.

Recent studies have found that obesity is associated with enlarged axillary LNs on screening mammograms secondary to fat expansion of the radiolucent LN hilum without accompanying enlargement of the nodal cortex as demonstrated in **Figure 1(b)** [10, 11]. There is marked variability in the degree of fatty node enlargement among obese women with similar body mass index (BMI), yet the clinical significance of this variability is unknown (**Figure 1**). Obesity-related fat deposition in other organs such as the liver, kidney, and bone marrow is associated with altered lipid metabolism that may lead to organ dysfunction, increased risk of malignancy, and poor cancer outcomes [12–16]. There are several proposed mechanisms for obesity-associated poor-prognosis cancer. Obesity-related adipose inflammation and dysregulated lipid metabolism can lead to increased secretion of inflammatory markers and adipokines that promote angiogenesis and tumor growth. Surplus local fat may be used as fuel by malignant cells and may additionally provide essential phospholipid building blocks required for cell membrane synthesis within proliferating tumors [17]. However, there has been very little research exploring fat deposition within LNs, the organelles of the immune and lymphatic system distributed throughout the body. We hypothesized that fat-infiltrated axillary LNs may be subject to similar adipose-induced dysfunction identified in other organs infiltrated by fat; and that the changes exerted by excess hilar fat may impact host resistance, potentially contributing to a higher risk of axillary metastases. Studies evaluating the significance of fat-expanded nodes may improve our understanding of mechanisms responsible for increased risk of node-positive, poor-prognosis breast cancer among obese women. Our study aimed to evaluate the relationship between axillary metastases and the size of fatty axillary nodes visualized on breast MRI and mammograms among obese women with invasive breast cancer.

**Figure 1.**
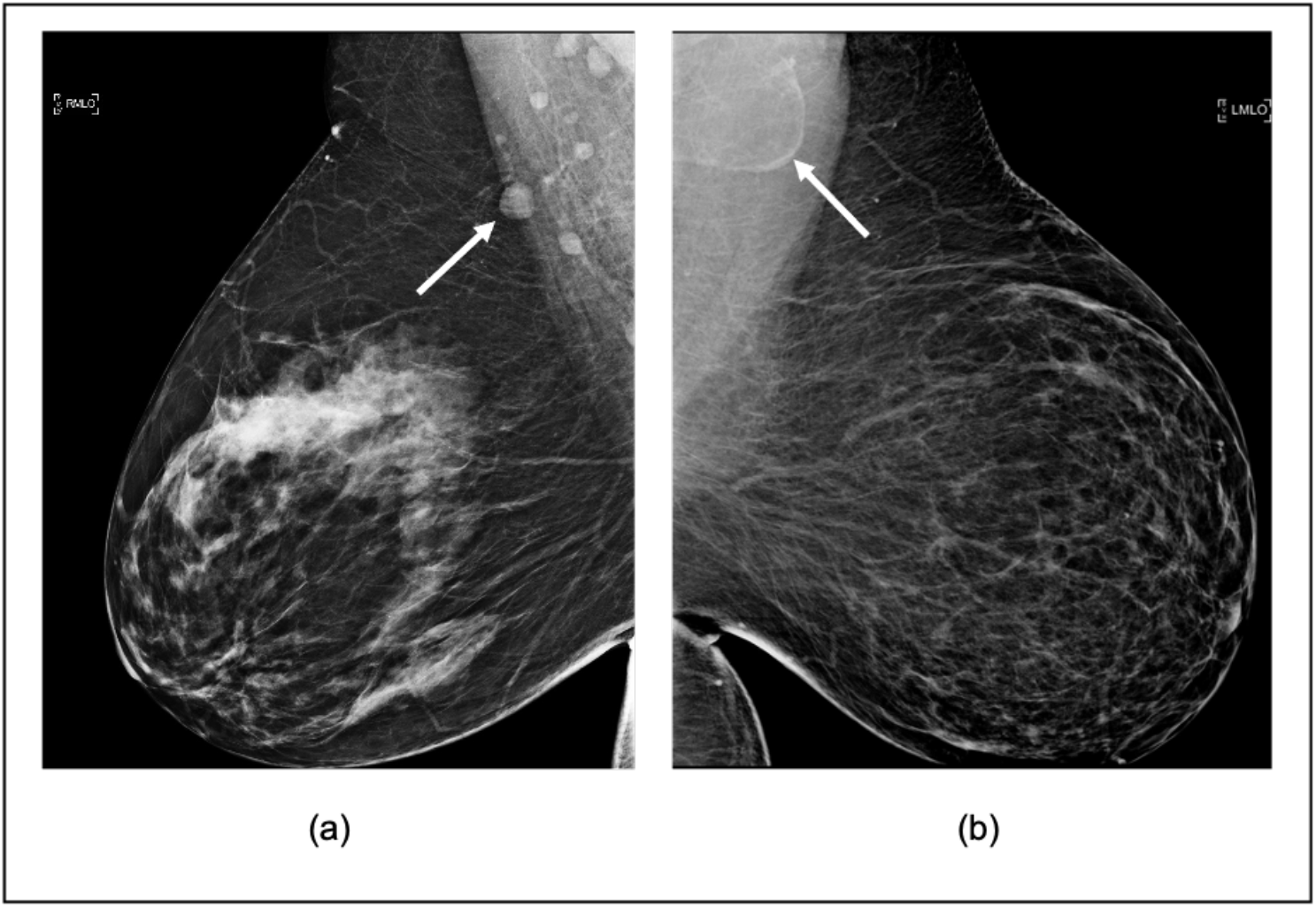
Variable axillary lymph node size and morphology on mammography. Obese women with variable fatty node morphology in the axilla on mediolateral oblique (MLO) views. **(a)** Normal axillary LNs measuring less than 1.5 cm in a woman (60’s) with BMI = 43.2. **(b)** fat-infiltrated LN measuring 4.2 cm in a woman (50’s) with BMI=45.8. Arrows point to the largest visible axillary lymph node.

## Materials and Methods

### Data Collection

This retrospective case-control study was approved by an institutional review board and was compliant with the Health Insurance Portability and Accountability Act. The dataset was collected from the Institutional Cancer Center Database identifying all obese women (BMI > 30) with histologically confirmed invasive breast cancer diagnosed between April 1, 2011 and March 1, 2020. An institutional review board exempted informed consent from these patients. Exclusion criteria included: imaging studies not available for review, pathology report not available for review, age greater than 89, history of recurrence, adenopathy secondary to malignancy other than breast cancer, isolated tumor cells on node histology, LVI status not available, or bilateral synchronous breast cancer with bilateral axillary metastases. Patients’ nodal status was determined according to the surgical pathology report or pre-operative LN biopsy histology report in patients who had neoadjuvant chemotherapy prior to surgery. All histologically confirmed node-positive patients that fulfilled the inclusion criteria were included in the final dataset. Node-negative patients were randomly selected from the same time period and were subjected to the same inclusion and exclusion criteria, in a ratio of approximately one control to two cases.

We collected the following patient and tumor characteristics from the electronic medical record as potential confounders: patient’s age at initial diagnosis, BMI, tumor size, tumor grade, estrogen receptor (ER), progesterone receptor (PR), and human epidermal growth factor receptor 2 (HER2) status, presence of lymphovascular invasion (LVI), and treatment with neoadjuvant chemotherapy or neoadjuvant endocrine therapy. For patients treated with neoadjuvant systemic therapy, tumor size on pre-treatment breast MRI was used in the analysis, while tumor size from surgical pathology reports was used for all other patients.

### Image Analysis

Axillary LNs were measured on breast MRI and mammograms by a breast radiologist with 18 years of experience. To assess the inter-observer agreement of LN measurements, a second breast radiologist with 19 years of experience independently measured LN size for 28% of the patients randomly selected from the original dataset. We used the largest LN visualized on breast MRI in the contralateral axilla for our primary analysis. The single largest LN within the axilla was chosen as the index node and measured along its greatest longitudinal axis in the axial or sagittal plane as shown in **Figure 2 and Figure 3**. Analysis of lymph node measurements visualized mammographically in the contralateral axilla is available in the Supplementary Material and measured as described in the prior study [10]. In order to avoid potential inclusion of morphologically normal nodes with micro-metastases occult to imaging in the ipsilateral axilla, we did not include measurements of ipsilateral axillary lymph nodes on MRI or mammography in the primary analysis, but they are available in the **Supplementary Materials**. Images were reviewed on Barco 3-megapixel MDCG-3221 monitors (Kortrijk, Belgium) with Philips PACS v. 3.6 (Philips Healthcare; Best, Netherlands).

**Figure 2.**
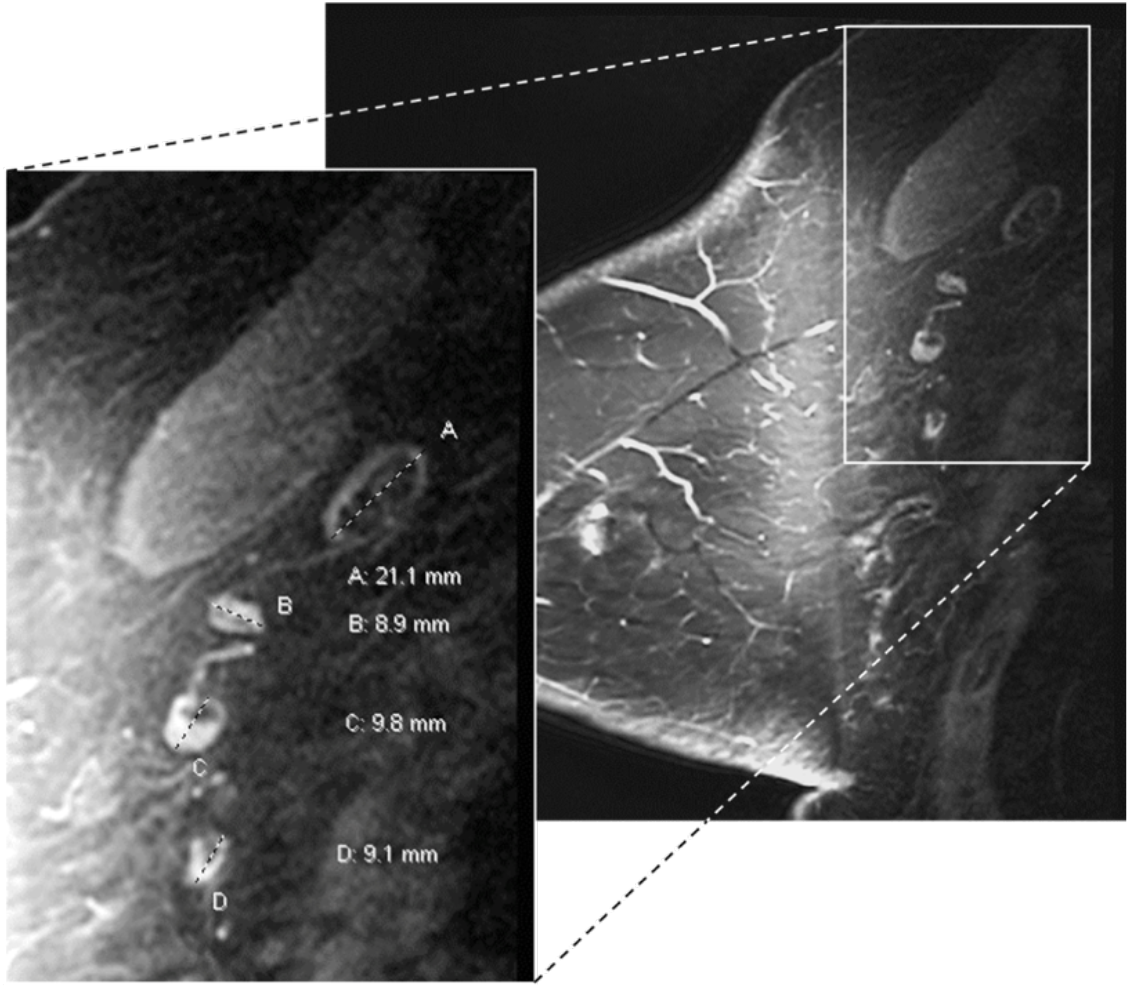
Variable axillary lymph node size and morphology on breast MRI. A female (60’s) with ER+ HER2-infiltrating ductal cancer of the left breast showing benign variable lymph node size and morphology in the right axilla contralateral to the known breast cancer. Contrast-enhanced fat-saturated sagittal MRI image of the right breast, and enlarged view of the right axilla demonstrate variable nodal size and morphology. Fat-enlarged node with expanded fatty hilum measures 21mm in length (A) while normal nodes with small fatty hila measure less than 10mm (B, C, D). The largest visible axillary node of 21 mm was chosen as the index node for the analysis in our study.

**Figure 3.**
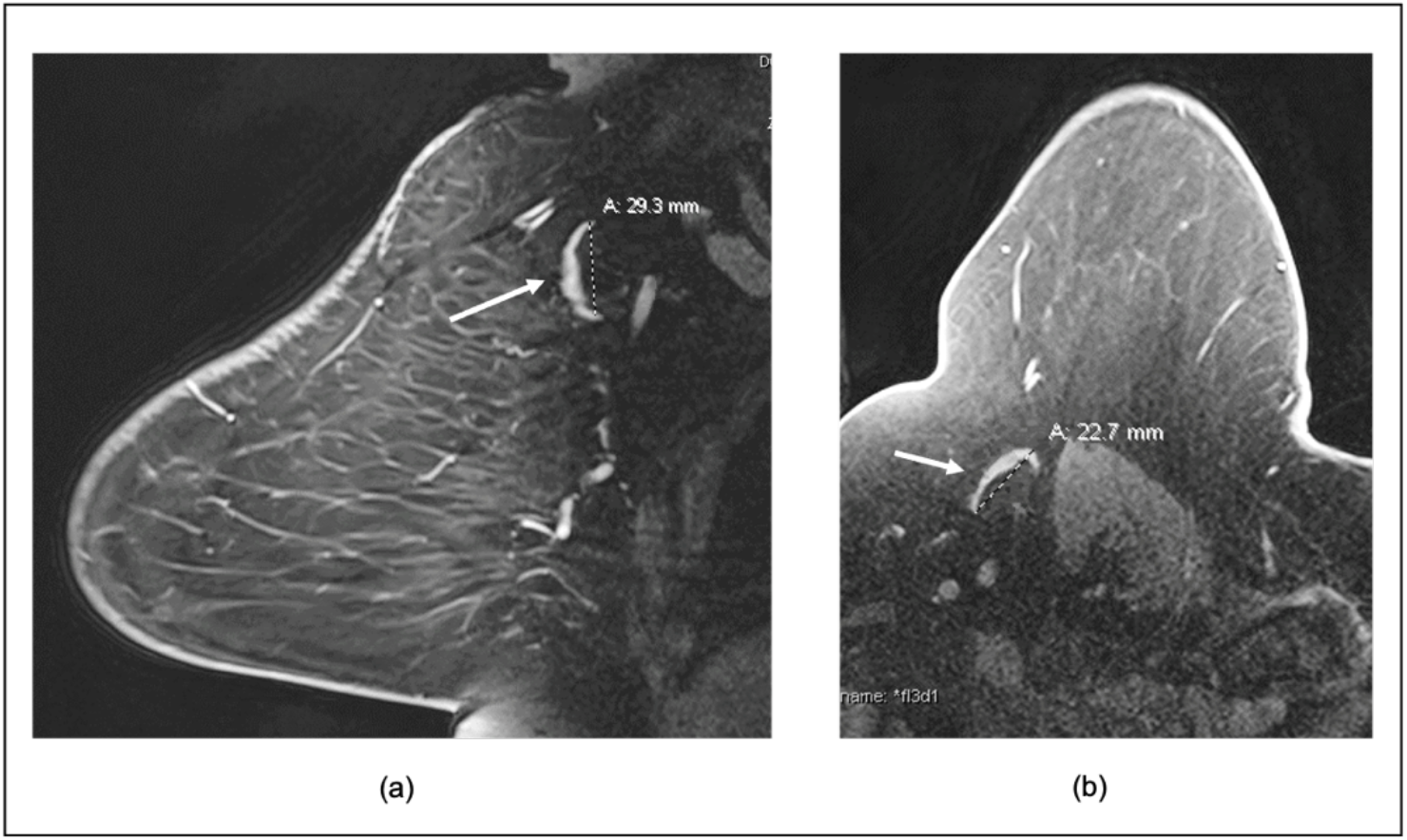
Axillary node measurements on breast MRI. A female (70’s) with left breast 18mm ER+HER2-node-positive invasive ductal cancer showing differences in the measurements of a single lymph node in the contralateral right axilla in the sagittal and axial plane. **(a)** Sagittal contrast enhanced fat-saturated breast MRI image through the contralateral right axilla demonstrates a fat-enlarged lymph node measuring 29mm in greatest dimension. **(b)** Axial contrast enhanced image through the same index node shows smaller greatest dimension of 23mm. We chose the largest measurement (in the sagittal or axial plane) for our study and in this case, we used 29mm for the analysis as identified in the sagittal plane from (a).

## Statistical Analysis

We conducted an independent sample t-test to compare the mean index LN size of the largest visible axillary node on breast MRI and mammograms. We further calculated the pairwise Pearson correlation between the index LN sizes measured on breast MRI and mammograms. Pearson correlation also was used to evaluate the inter-observer reliability of LN measurements between two radiologists. We examined the association between node-positive breast cancer and fat-enlarged axillary LN size using multivariate logistic regression to adjust for covariates of interest. We conducted separate analyses on LNs visualized on breast MRI and mammograms. In each analysis, LN measurements were categorized into quartiles containing equal numbers of cases. The quartile with the smallest LN sizes was used as the reference group in the regression analyses. Adjusted odds ratios (OR) with 95% confidence intervals (CI) were calculated for LN size quartiles. All statistical analyses were performed using R software (version 4.0.3; RStudio, Boston, Mass). Receiver operative characteristic (ROC) curves with 5-fold cross-validation were generated to evaluate the ability to discriminate node-positive from node-negative patients using LN size. ROC curves were obtained with LN size alone and combining LN size with other clinical variables including age, BMI, tumor size, tumor grade, LVI, and neoadjuvant systemic therapy. ROC curves and cross-validation were done with Python programming language (Version 3.7.1).

## Results

A total of 355 patients with node-positive breast cancer and BMI > 30, diagnosed between April 1, 2011, and March 1, 2020, were identified by the Institutional Cancer Center Database. 71 patients were excluded, and the reasons are shown in **Figure 4**. The remaining 284 node-positive cases were combined with 147 random-selected node-negative controls to form our final dataset of 431 patients.

**Figure 4.**
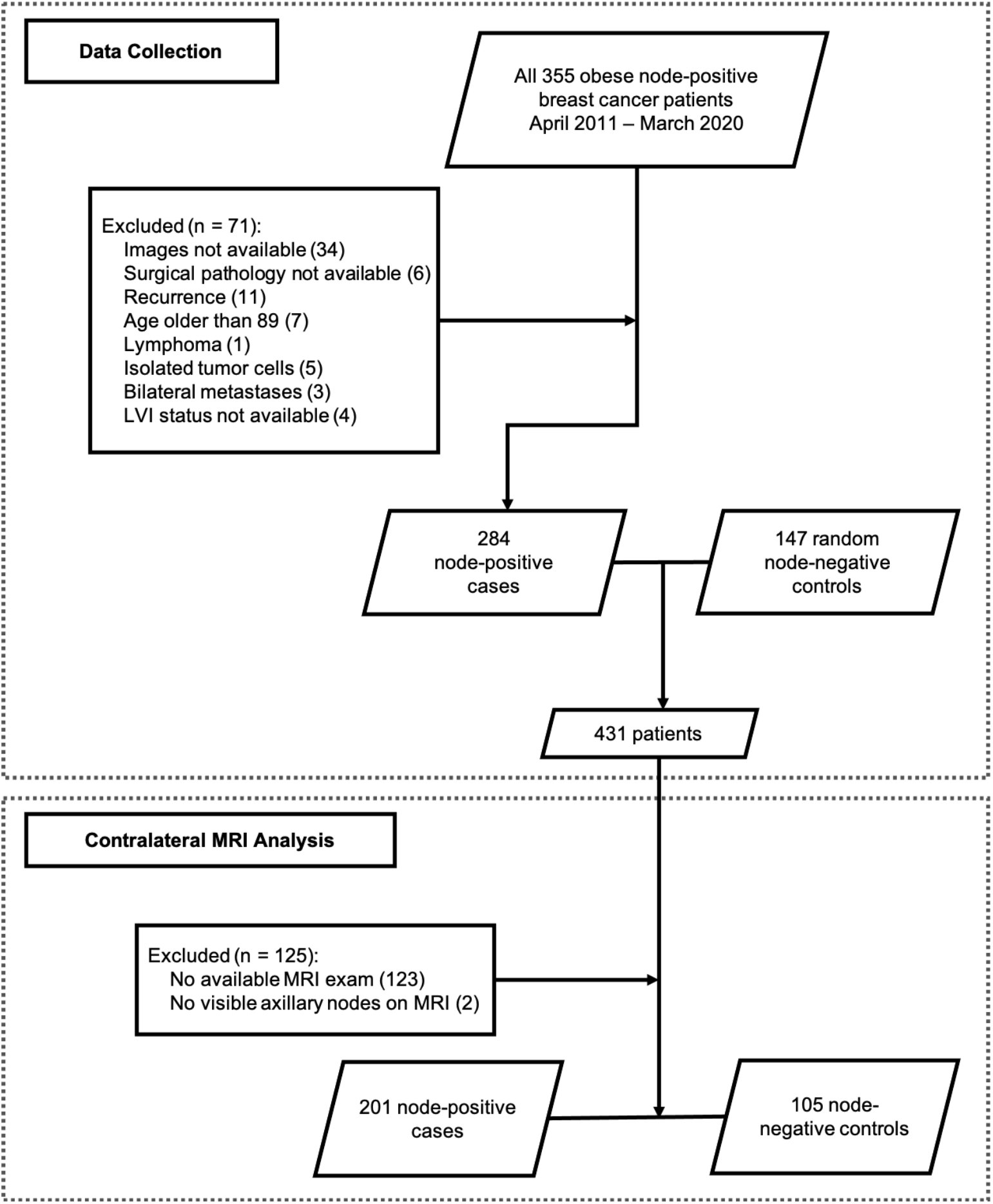
Flowchart of data collection for primary analysis evaluating LN size on breast MRI identified in the axilla contralateral to the known breast cancer.

We observed that LN sizes visualized on mammography were significantly smaller than those identified on breast MRI with mean size of 19.70 ± 7.42 mm on mammography, and 25.54 ± 7.29 on breast MRI (p < 0.001). Despite difference in mean size, LN measurements obtained mammographically and on breast MRI were positively correlated (r = 0.66, p < 0.001) (**Figure 5 and Figure 6**).

**Figure 5.**
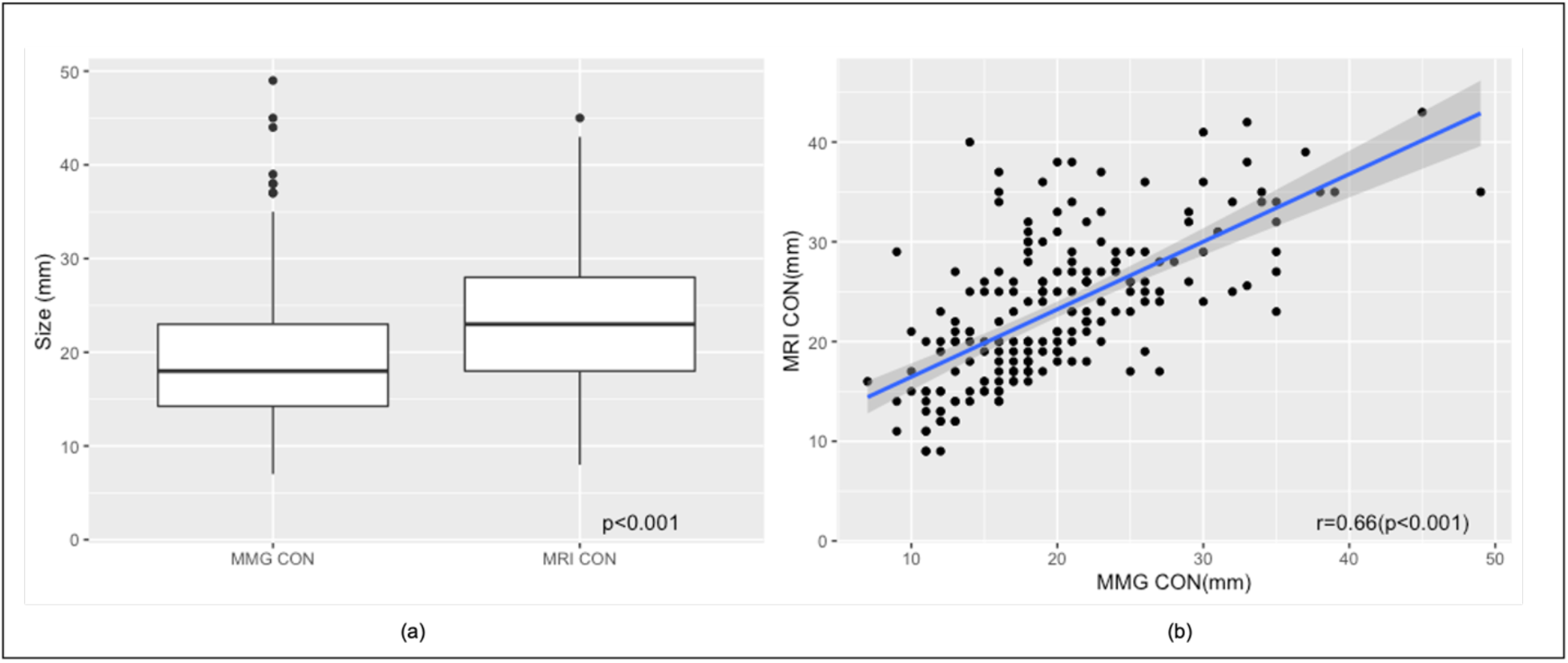
Correlation of LN measurements on mammogram and breast MRI. **a)** Distribution of axillary LN sizes measured on contralateral mammograms and breast MRI. **b)** Despite difference in mean size, LN measurements obtained mammographically and on breast MRI were positively correlated (r = 0.66, p < 0.001). Scatterplot of LN size on mammogram and breast MRI with fitted regression line (blue) and 95% CI (shaded) showing good correlation. MMG = mammogram, CON = contralateral.

**Figure 6.**
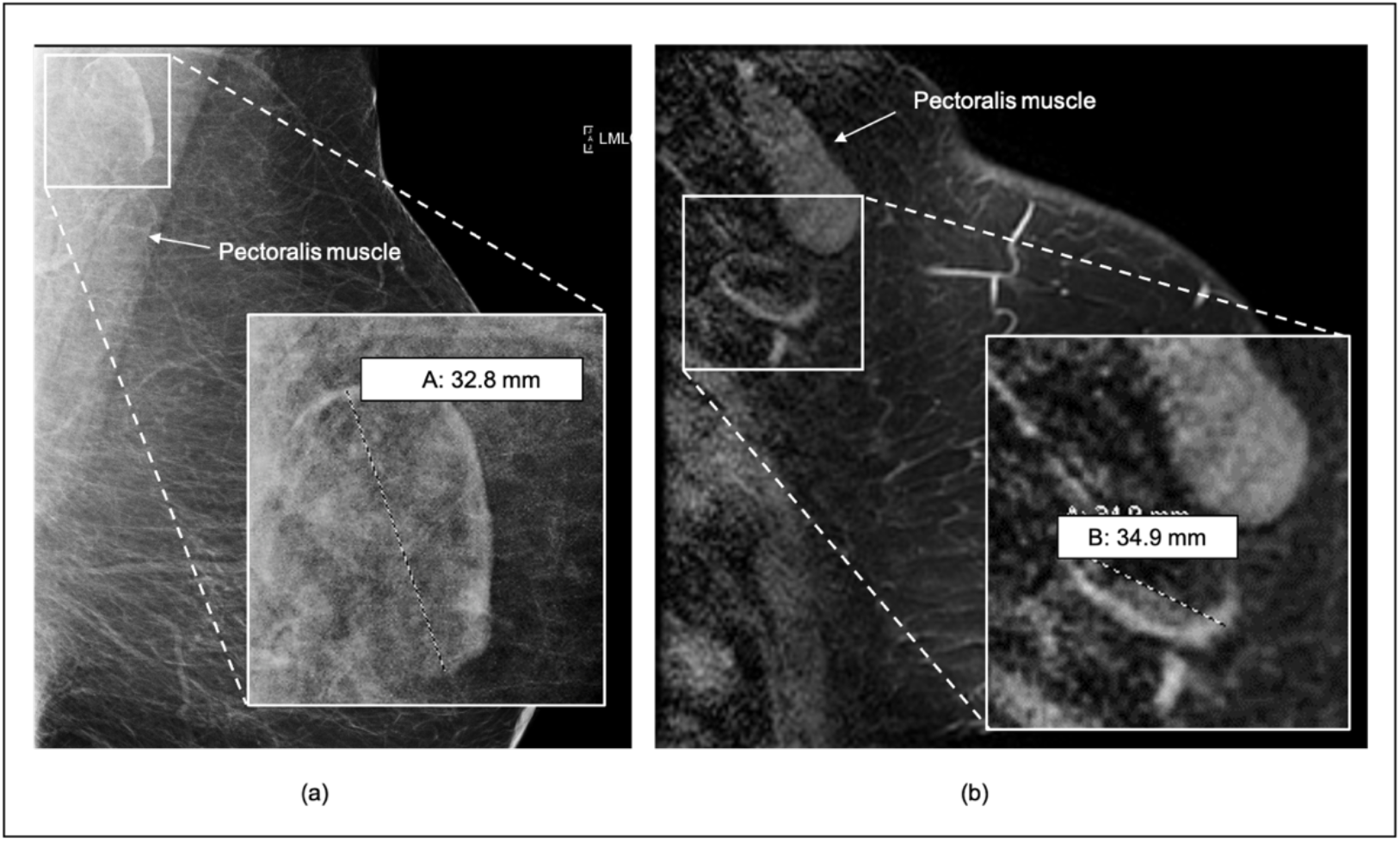
Comparison of LN measurements on mammogram and breast MRI. Right breast of a female (50’s) with ER+HER2+ invasive ductal carcinoma demonstrating contralateral fat-expanded node on mammography compared to breast MRI. **(a)** Left MLO digital mammogram shows a fat-expanded lymph node in the contralateral axilla measuring 33 mm in largest dimension, **(b)** sagittal fat-saturated enhanced breast MRI of the same patient demonstrates a slightly larger size of the same lymph node measuring 35 mm in length on MRI.

LNs were almost always identified on breast MRI consistent with a larger field of view that usually allows comprehensive evaluation of the entire axilla and demonstrates a larger number of axillary nodes. Based on the second radiologist’s assessment of 28% of patients, the measurement of contralateral LNs on MRI showed strong inter-observer agreement with a Pearson correlation coefficient of 0.67 (p < 0.001). Our primary analysis consists of LN measurements in the contralateral axilla on breast MRI. The following patients were excluded: 123 patients without breast MRI available for review, and 2 patients without visible contralateral axillary lymph nodes on breast MRI, resulting in a total of 306 patients (201 cases and 105 controls) included in the primary analysis of MRI-visualized contralateral axillary lymph nodes (**Figure 4**). The demographics and clinical characteristics of the patients in the primary analysis of MRI-visualized contralateral axillary lymph nodes are shown in **Table 1**. The patient demographics and analysis results of contralateral mammographic LNs and of ipsilateral MRI and mammographic LNs can be found in the **Supplementary Materials**.

**Table 1.** Characteristics of patients used for contralateral MRI analysis (n = 306).

|  | **Node Negative** | **Node Positive** | **P-value** |
| --- | --- | --- | --- |
| **N (%)** | 105 (34.3) | 201 (65.7) |  |
| **Age (years, SD)** | 60.67 (10.1) | 57.73 (10.6) | 0.020 |
| **BMI (SD)** | 36.33 (5.8) | 35.86 (4.9) | 0.458 |
| **Tumor size (mm, SD)** | 25.83 (17.1) | 37.52 (23.8) | <0.001 |
| **Tumor grade (%)** |  |  | 0.032 |
| **1** | 23 (21.9) | 22 (10.9) |  |
| **2** | 47 (44.8) | 96 (47.8) |  |
| **3** | 35 (33.3) | 83 (41.3) |  |
| **Molecular Subtypes (%)** |  |  | 0.022 |
| **ER+ HER2-** | 81 (77.1) | 147 (73.1) |  |
| **HER2+** | 9 (8.6) | 38 (18.9) |  |
| **TNBC** | 15 (14.3) | 16 (8.0) |  |
| **NAC or NAE (%)** | 5 (4.8) | 56 (27.9) | <0.001 |
| **LVI (%)** | 22 (21.0) | 117 (58.2) | < 0.001 |
| **MRI CON LN Size (mm, SD)** | 19.91 (6.93) | 25.43 (7.0) | <0.001 |
| **[8, 18] (%)** | 54 (51.4) | 31 (15.4) |  |
| **(18, 23] (%)** | 23 (21.9) | 49 (24.4) |  |
| **(23, 28] (%)** | 15 (14.2) | 59 (29.4) |  |
| **(28, 45] (%)** | 13 (12.4) | 62 (30.9) |  |

<sup>a</sup> BMI = body mass index, ER = estrogen receptor, HER2 = human epidermal growth factor receptor 2, TNBC = triple negative breast cancer, NAC = neoadjuvant chemotherapy, NAE = neoadjuvant endocrine, LVI = lymphovascular invasion, CON = contralateral, LN = lymph node

### Association between fatty nodes and nodal status

Compared to the baseline LN size (< 18mm), statistically significant positive associations were observed between node-positive breast cancer and larger fatty nodes in the contralateral axilla on breast MRI adjusting for age, BMI, tumor size, tumor grade, tumor molecular subtype, and LVI. Specifically, contralateral LN size greater than 28 mm (4^th^ quartile) had an estimated odds ratio of 9.70 compared to the first quartile (95% CI: 4.26, 23.50; p-value < 0.001), and the estimated odds ratio increased with nodal size. We observed a similar positive association between axillary LN size and node-positive breast cancer with contralateral mammographic LN, and ipsilateral mammographic and breast MRI-visualized LNs as shown in Supplementary tables 2 and 3. Of note, contralateral LN size significantly correlated with ipsilateral LN size, assuring the validity of using contralateral LNs for the primary analysis (**Supplementary Materials Figure 1**).

### Association between fatty LN size and nodal status in patients without LVI

LVI remained strongly significantly associated with positive nodal status, and we observed larger LN sizes in patients with LVI compared to those without (p < 0.001). From our dataset, 167 patients had no LVI, of whom 84 (50.3%) had nodal metastasis. Our analysis showed that for patients without LVI, increased LN size was associated with an increased likelihood of nodal metastasis, adjusting for other collected variables. The association between contralateral LN size and nodal metastasis in patients without LVI is shown in **Table 3**.

**Table 2.**
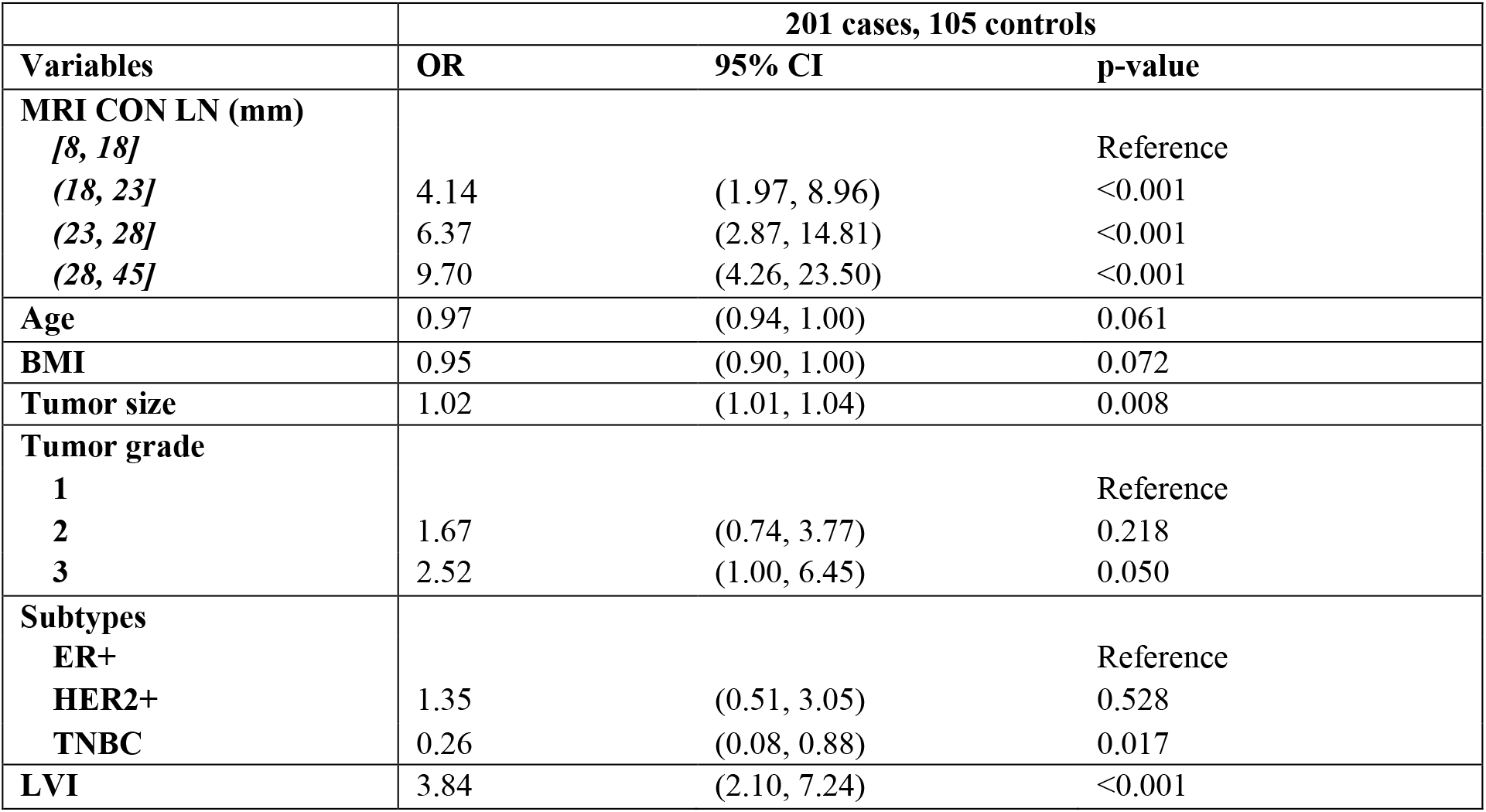
**Multivariate logistic regression analysis of association between index LN size on contralateral MRI adjusting for potential confounders (N = 306)**.

**Table 3.** Association between LN size and nodal status in the subgroup without the presence of LVI.

| MRI CON LN (mm) | No LVI (84 cases, 83 controls) |  |  |
| --- | --- | --- | --- |
|  | OR | 95% CI | p-value |
| [8, 17] |  |  | Reference |
| (17, 21] | 5.26 | (1.83, 16.28) | <0.003 |
| (21, 27] | 9.23 | (3.32, 28.22) | <0.001 |
| (27, 40] | 17.46 | (5.76, 60.20) | <0.001 |

<sup>a</sup> The analyses were adjusted for potential confounders including age, BMI, tumor size and grade, molecular subtypes, and the presence of LVI (results not shown in the table). CON = contralateral.

### Predictability of axillary node metastases by size of fat-enlarged contralateral axillary LNs

**Figure 7** shows the ROC curves for axillary metastasis using LN size alone and combining LN size with patient and tumor characteristics, including age, BMI, tumor size, tumor grade, molecular subtype, presence of LVI, and neoadjuvant systemic therapy. The prediction with LN size alone reached an area under the ROC curve (AUC) of 0.72 ± 0.05, while including LN size with other variables as predictors improved the AUC to 0.80 (**Figure 7**). The ROC curves of contralateral mammographic nodes and ipsilateral nodes from both mammograms and MRI are shown in **Supplementary Materials S3**.

## Discussion

We found that enlarged fat expanded axillary lymph nodes were strongly associated with node-positive breast cancer in obese women. The area under the ROC curve for the association between axillary metastases and enlarged fatty nodes in the contralateral axilla on breast MRI was 72%, and this increased to 80% when combined with other clinical variables. The mean size of MRI-detected nodes was significantly larger than mammographically-detected nodes, possibly reflecting differences in LN size related to patient position, degree of compression, multiple imaging planes, and improved resolution of nodes on MRI compared to mammography. Contralateral nodes identified by MRI were chosen for the primary analysis in our study in order to avoid the potential inclusion of morphologically normal nodes with micro-metastases occult to imaging in the ipsilateral axilla, and to permit the largest number of visible axillary nodes for analysis. Interestingly, both ipsilateral and contralateral enlarged fatty lymph nodes visualized on breast MRI and mammography were independently associated with nodal metastases as indicated in the supplementary material.

**Figure 7.**
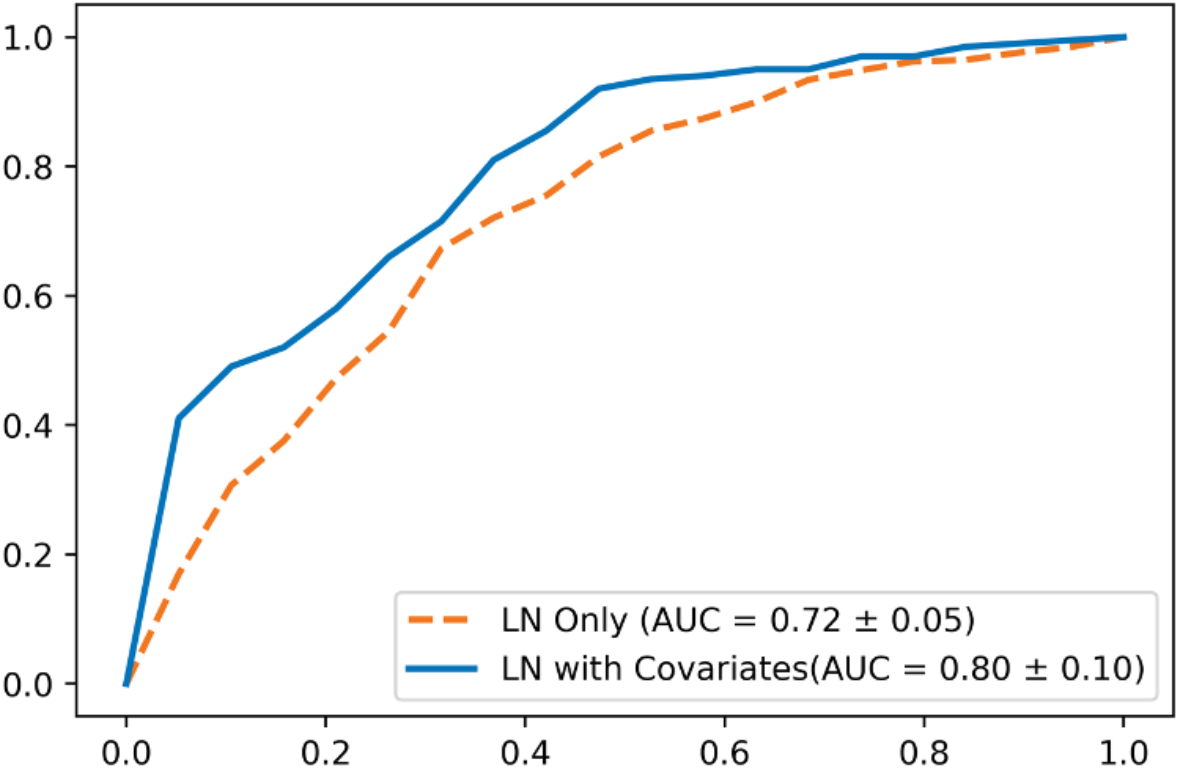
Performance of contralateral fat-enlarged node size for predicting axillary metastases. Mean ROC curves of node positive breast cancer prediction using axillary LN size from contralateral MRI with 5-fold cross-validation. The orange dash line indicates prediction of axillary metastases using contralateral MRI LN size alone. The blue solid line indicates prediction of axillary metastases using contralateral LN size combined with other variables.

Studies have underscored the need to refine BMI into features of body composition to clarify the association between obesity and poor prognosis cancer. Ectopic fat deposition within organs has been shown to represent a better predictor of adverse health outcomes, increased malignancy risk, and poor cancer outcomes compared to BMI or subcutaneous fat [14, 18–22]. Our study demonstrated that fatty nodes were strongly associated with axillary metastases in obese patients, while BMI was not. Although there is a lack of research evaluating LN fat deposition in humans, findings in obese mice demonstrate impaired immune function and decreased lymphatic transport associated with fat deposition within LNs and lymphatics [23]. Within the breast, studies evaluating lipid function and metabolism have found associations between altered fat composition and breast cancer. Changes in fatty acid oxidation are associated with increased breast cancer proliferation and poor outcomes [17, 24]. High expression of Spot14, a requisite gene for fatty acid synthesis, is associated with decreased survival in breast cancer patients [25]. Differences in fatty acid fractions have been observed in breast adipose tissue of postmenopausal women with breast cancer compared to women without breast cancer, independent of BMI [26]. Additionally, MR spectroscopy has identified lipid dysregulation within breast tissue of women with BRCA gene mutations [27].

LVI indicates the presence of tumor cells within the peri-tumoral vascular or lymphatic channels and is a strong prognostic marker for axillary metastases. LVI is therefore included as a predictive feature in models designed to assess the likelihood of positive sentinel nodes as well as positive non-sentinel node axillary metastases [28, 29]. In our study, half of the patients without LVI developed nodal metastasis prompting us to perform additional analysis to better understand the relationship between fat-enlarged nodes and nodal metastasis in patients without LVI. Our findings demonstrated a strong association between fatty node size and node-positive status among patients without LVI. A proposed mechanism for this interesting observation could be that LVI and fat-enlarged nodes operate via divergent mechanisms in the invasion-metastasis cascade. The status of the axilla in breast cancer patients reflects the interaction between tumor aggressiveness and host resistance. LVI is most commonly seen with larger tumor size and higher histologic grade, suggesting that aggressive tumor characteristics influence the predictive nature of LVI [30]. In contrast, one possible explanation for our results showing that fat-enlarged nodes were strongly associated with axillary metastases in women without LVI is that fatty nodes may reflect features of host resistance rather than the metastatic potential of the tumor.

There are several potential mechanisms by which excess hilar fat may contribute to an increased likelihood of nodal metastases. Structurally, excess hilar adipose may compress traversing arteries, veins, and efferent lymphatics, potentially compromising nodal function by decreasing vascular flow and lymphatic clearance of isolated tumor cells. A similar mechanism of fat compression in obesity has been described in the kidney, an organ that is structurally very similar to lymph nodes, wherein excess renal sinus fat compression of vessels has been linked to renal dysfunction and hypertension [31]. Increased hilar fat may additionally support the establishment of axillary metastases via mechanisms related to changes in the LN adipose microenvironment as described in other ectopic fat depots. These include chronic low grade inflammation and alterations in lipid metabolism within obese adipose tissue that support the establishment and proliferation of malignant tumors [7, 32–36].

Our study is limited by its retrospective nature. Mammograms and breast MRI were obtained in different imaging centers; however, patients were referred to a single academic institution for breast cancer treatment, which may limit generalizability. Pre-operative breast MRI was not available for all patients, was more likely to be obtained in younger women, and was furthermore driven by surgeon preference which could have introduced bias. Despite these limitations, our study indicates strong agreement of LN measurements on breast MRI between two independent breast imagers. While fatty nodes represent a benign variant relative to metastatic nodes, our findings suggest that enlarged fat-expanded axillary lymph nodes may represent an imaging biomarker of axillary metastases in obese women. Future studies are needed to confirm our findings and to improve our understanding of mechanisms responsible for the poor prognosis of breast cancer in obese women.

## Data Availability

The data that support the findings of this study are available from the corresponding author, S.H., upon reasonable request.

