## Supplementary Materials for "Fat-enlarged Axillary Lymph Nodes are Associated with Node-Positive Breast Cancer in Obese Patients"

One Medical Center Drive, HB 7261

Lebanon, NH 03756

### Supplementary Materials

#### S1. Analysis Results for Contralateral Mammographic Axillary LN

We conducted an analysis of LN size on contralateral mammograms, adjusting for collected covariates of interest. From 431 patients, we excluded 21 patients with no available contralateral mammogram for review and 132 with no visible axillary LNs in the contralateral axilla. The characteristics of the remaining patients are shown in **Table 1**. We observed a significant association between axillary LN size on contralateral mammograms and nodal metastasis with the results shown in **Table 2**.

**Table 1. Characteristics of patients with mammographic LN measurements in the contralateral axilla (n = 278).**

| | **Node Negative** | **Node Positive** | **P-value** |
|---|---|---|---|
| **N (%)** | 106 (38.1) | 172 (61.9) | |
| **Age (years, SD)** | 61.46 (9.3) | 60.55 (11.0) | 0.479 |
| **BMI (SD)** | 35.97 (5.5) | 36.56 (5.8) | 0.400 |
| **Tumor size (mm, SD)** | 25.01 (17.0) | 6.05 (23.0) | <0.001 |
| **Tumor grade (%)** | | | 0.014 |
| ***1*** | 24 (22.6) | 19 (11.1) | |
| ***2*** | 52 (49.1) | 83 (48.2) | |
| ***3*** | 30 (28.3) | 70 (40.7) | |
| **Molecular Subtypes (%)** | | | 0.018 |
| ***ER+ HER2-*** | 84 (79.3) | 129 (75.0) | |
| ***HER2+*** | 8 (7.6) | 31 (18.0) | |
| ***TNBC*** | 14 (13.2) | 12 (7.0) | |
| **NAC or NAE (%)** | 3 (2.8) | 39 (22.7) | <0.001 |
| **LVI (%)** | 22 (21.0) | 117 (58.2) | < 0.001 |
| **MMG CON LN Size (mm, SD)[1]** | 16.22 (5.5) | 21.84 (7.5) | <0.001 |
| ***[8, 18] (%)*** | 43 (40.6) | 27 (15.7) | |
| ***(18, 23] (%)*** | 38 (35.9) | 34 (19.8) | |
| ***(23, 28] (%)*** | 16 (15.1) | 54 (31.4) | |
| ***(28, 45] (%)*** | 9 (8.49) | 57 (33.1) | |

[a] Threshold of LN size categories were chosen based on quartiles of LN size of 278 patients in the sub-dataset. BMI = body mass index, ER = estrogen receptor, HER2 = human epidermal growth factor receptor 2, TNBC = triple negative breast cancer, NAC = neoadjuvant chemotherapy, NAE = neoadjuvant endocrine, LVI = lymphovascular invasion, MMG = mammogram, CON = contralateral

**Table 2. Multivariate logistic regression analysis of the association between nodal metastases and LN size in the contralateral axilla on mammography.**

| | **172 cases, 106 controls** | | |
|---|---|---|---|
| **MMG CON LN (mm)** | **OR** | **95% CI** | **p-value** |
| ***[7.0, 14.2]]*** | | | Reference |
| ***(14.2, 18.0]*** | 1.07 | (0.49, 2.33) | 0.87 |
| ***(18.0, 23.0]*** | 4.67 | (2.07, 10.97) | <0.001 |
| ***(23.0, 49.0]*** | 7.71 | (3.05, 21.22) | <0.001 |
| **Age** | 1.00 | (0.97, 1.04) | 0.791 |
| **BMI** | 1.00 | (0.96, 1.06) | 0.804 |
| **Tumor size** | 1.02 | (1.00, 1.04) | 0.015 |
| **Tumor grade** | | | |
| **1** | | | Reference |
| **2** | 1.41 | (0.61 3.27) | 0.418 |
| **3** | 2.57 | (0.98, 6.88) | 0.056 |
| **Subtypes** | | | |
| **ER+** | | | Reference |
| **HER2+** | 2.09 | (0.81, 5.83) | 0.139 |
| **TNBC** | 0.22 | (0.07, 0.70) | 0.011 |
| **LVI** | 4.56 | (2.45, 8.76) | <0.001 |

[a] BMI = body mass index, ER = estrogen receptor, HER2 = human epidermal growth factor receptor 2, TNBC = triple negative breast cancer, NAC = neoadjuvant chemotherapy, NAE = neoadjuvant endocrine, LVI = lymphovascular invasion, MMG = mammogram, CON = contralateral

#### S2. Association between Ipsilateral Axillary LN Size and axillary metastases

We calculated the Pearson correlation for ipsilateral and contralateral LN sizes measured on breast MRI of 105 node negative patients. **Supplementary Figure 1** showed strong significant correlation between LN sizes in both axilla supporting the validity of using contralateral LN size for our primary analysis. In addition to contralateral nodes, we also conducted analyses of LN size within the ipsilateral axilla. In patients with low axillary tumor burden (< 3 metastatic nodes), we measured the largest visible, morphologically normal, non-metastatic LN within the ipsilateral axilla.

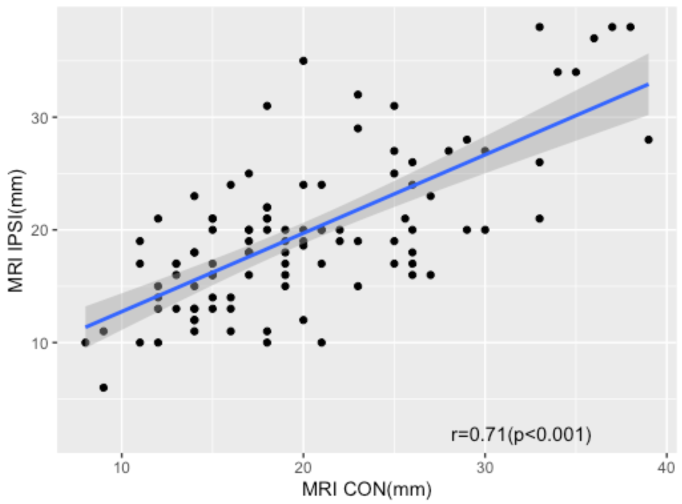


**Figure 1. Correlation between ipsilateral and contralateral LN.** LN measurement from contralateral vs ipsilateral axilla in node-negative patients demonstrated a strong positive correlation between node sizes in both axilla with a Pearson correlation coefficient of 0.71 ($p<0.001$). IPSI = ipsilateral, CON = contralateral.

We conducted multivariate logistic regression to examine the association between ipsilateral LN size and node-positive breast cancer on both breast MRI and mammograms, as described in the methods section. For each analysis, we excluded the following from 431 patients in the dataset: ipsilateral breast MRI: no studies available for review (118), greater than three metastatic nodes (32), no visible LN (3); ipsilateral mammogram: no available images for review (16), greater than three metastatic nodes (17), no visible LNs (134). As observed with contralateral LN, we similarly observed a significant association between axillary LN size and nodal metastasis in the ipsilateral axilla. The number of cases and controls and the analysis results in each sub-analysis are shown in **Table 3**.

**Table 3. Multivariate logistic regression analysis of association between ipsilateral index LN size and nodal status.**

a) Ipsilateral Breast MRI

| | 172 cases, 106 controls | | |
|---|---|---|---|
| **MRI IPSI LN (mm)** | **OR** | **95% CI** | **p-value** |
| ***[6.0, 18.0]*** | | | Reference |
| ***(18.0, 22.0]*** | 2.89 | (1.35, 6.38) | 0.007 |
| ***(22.0, 26.8]*** | 11.02 | (4.56, 28.89) | <0.001 |
| ***(26.8, 52.0]*** | 5.12 | (2.28, 11.97) | <0.001 |

b) Ipsilateral Mammogram

| | 157 cases, 107 controls | | |
|---|---|---|---|
| **MMG IPSI LN (mm)** | **OR** | **95% CI** | **p-value** |
| ***[0.0, 15.0]*** | | | Reference |
| ***(15.0, 18.0]*** | 1.28 | (0.56, 2.89) | 0.553 |
| ***(18.0, 24.0]*** | 3.71 | (1.69, 8.41) | 0.001 |
| ***(24.0, 50.0]*** | 8.69 | (3.45, 24.04) | <0.001 |

[a] Threshold of LN size categories were chosen based on quartiles of LN size in each sub-dataset. The analyses were adjusted by age, BMI, tumor size, tumor grade, molecular subtype, and presence of LVI (results not shown). MMG = mammogram, IPSI = ipsilateral, CON = contralateral, OR = odds ratio, CI = confidence interval.

#### S3. Predictability of LN Size from Contralateral Mammograms and in Ipsilateral Axilla

We plotted ROC curves for prediction of axillary metastases from LN size within the contralateral axilla on mammography as well LN size from the ipsilateral axilla on both breast MRI and mammography. Our results showed that LN node size alone can predict nodal metastasis with an AUC of 0.72 – 0.75, and when combined with patients' clinical information, the AUC improved it to 0.78 – 0.79 (**Supplementary Figure 2**).

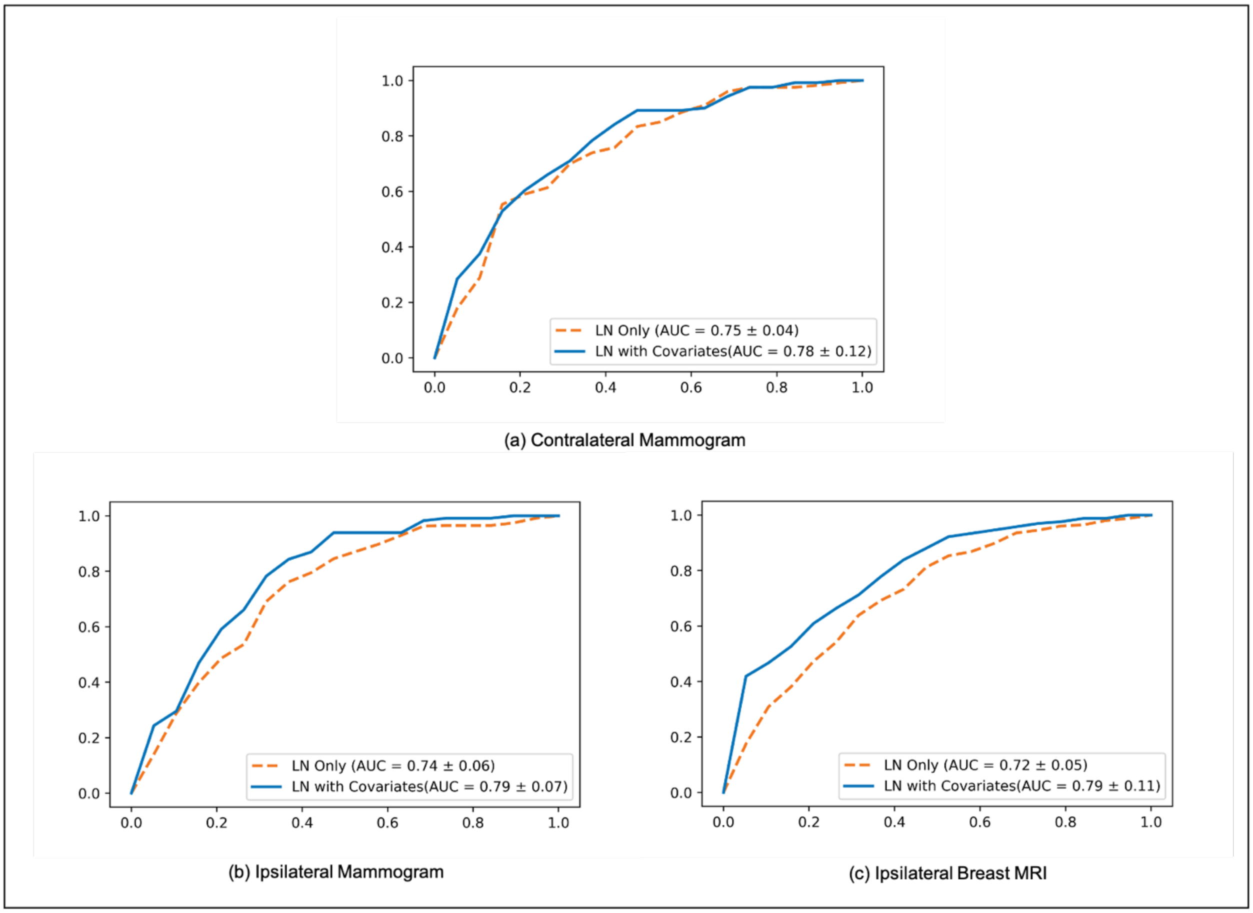


**Figure 2.** Mean ROC curves of node-positive breast cancer prediction with 5-fold cross-validation using LN size measured from (a) contralateral mammogram, (b) ipsilateral mammogram, and (c) ipsilateral breast MRI. Orange dash lines indicate prediction using contralateral LN size alone. Blue solid lines indicate prediction using LN size combined with other variables.
